# Chopping the tail: how preventing superspreading can help to maintain COVID-19 control

**DOI:** 10.1101/2020.06.30.20143115

**Authors:** Morgan P. Kain, Marissa L. Childs, Alexander D. Becker, Erin A. Mordecai

**Author notes:** Denotes equal authorship (Morgan P. Kain, Marissa L. Childs).

## Abstract

Disease transmission is notoriously heterogeneous, and SARS-CoV-2 is no exception. A skewed distribution where few individuals or events are responsible for the majority of transmission can result in explosive, superspreading events, which produce rapid and volatile epidemic dynamics, especially early or late in epidemics. Anticipating and preventing superspreading events can produce large reductions in overall transmission rates. Here, we present a compartmental (SEIR) epidemiological model framework for estimating transmission parameters from multiple imperfectly observed data streams, including reported cases, deaths, and mobile phone-based mobility that incorporates individual-level heterogeneity in transmission using previous estimates for SARS-CoV-1 and SARS-CoV-2. We parameterize the model for COVID-19 epidemic dynamics by estimating a time-varying transmission rate that incorporates the impact of non-pharmaceutical intervention strategies that change over time, in five epidemiologically distinct settings—Los Angeles and Santa Clara Counties, California; Seattle (King County), Washington; Atlanta (Dekalb and Fulton Counties), Georgia; and Miami (Miami-Dade County), Florida. We find the effective reproduction number ℛ_*E*_ dropped below 1 rapidly following social distancing orders in mid-March, 2020 and remained there into June in Santa Clara County and Seattle, but climbed above 1 in late May in Los Angeles, Miami, and Atlanta, and has trended upward in all locations since April. With the fitted model, we ask: how does truncating the tail of the individual-level transmission rate distribution affect epidemic dynamics and control? We find interventions that truncate the transmission rate distribution while partially relaxing social distancing are broadly effective, with impacts on epidemic growth on par with the strongest population-wide social distancing observed in April, 2020. Given that social distancing interventions will be needed to maintain epidemic control until a vaccine becomes widely available, “chopping off the tail” to reduce the probability of superspreading events presents a promising option to alleviate the need for extreme general social distancing.

## Introduction

In the face of emerging epidemics with limited pharmaceutical options for treatment and prevention of infection, non-pharmaceutical interventions such as social distancing are critical for slowing epidemic growth. Shelter-in-place and other social distancing orders have helped to slow the pace of the COVID-19 pandemic, reducing the effective reproduction number ℛ_*E*_—or the number of secondary infections produced by each infected person—to one or below in most places. In doing so, social distancing has effectively kept most regional healthcare systems operating under maximum capacity. However, after only a few weeks of declining numbers of daily cases due to an ℛ_*E*_ at or below one, most state and county governments in the United States have begun relaxing social distancing orders, citing their major economic impacts. In order to avoid epidemic resurgence, it is vitally important that governments employ long-term strategies that maintain epidemic control as economic reopening commences.

One obstacle to designing effective long-term strategies is a notoriously heterogeneous transmission process. It is now widely recognized that the minority of infections generate the majority of secondary cases, leading to the so-called 20-80 rule in epidemiology (the rule-of-thumb that 20% of people generate 80% of cases) ^1^. Work on SARS-CoV-1, measles, and other respiratory viruses suggests that this skew in secondary cases is even larger ^2^. This heterogeneity gives rise to events in which a single infected person transmits a disease to dozens or hundreds of people—called superspreading events—which have played an important role in the COVID-19 pandemic ^3,4,5,6,7^. Indeed, the frequency of asymptomatic and presymptomatic transmission, potential disconnect between infection and clinical presentation ^8^, and potential transmission via direct contact, aerosols, and surfaces ^9,10^ are all features of SARS-CoV-2 that tend to promote superspreading. As local and national governments search for viable exit strategies from shelter-in-place, a critical question is how effective curtailing superspreading events could be in controlling epidemic spread.

Practically, one strategy to help prevent superspreading is to prohibit medium to large indoor gatherings such as exercise classes, sporting events, concerts, and weddings for an extended period after allowing smaller and lower-risk activities to resume. From a modeling standpoint, predicting the effects of this straightforward intervention is difficult for two reasons: 1) local epidemiological dynamics are changing with evolving intervention strategies; and 2) information may not be available to parameterize detailed models of disease spread through heterogeneous populations. Despite these difficulties, it is important to consider some individual-level heterogeneity in transmission because model analyses of mean transmission rates alone may over-estimate the effectiveness of interventions, overlook potentially effective interventions that act on the heterogeneity within populations, overlook potentially explosive resurgences, and poorly predict the final epidemic size ^2,7^.

Studies of superspreading often empirically estimate secondary case distributions from recorded transmission chains and/or using branching process models ^2,5,6,11^. These studies estimate a dispersion parameter, *k*, that describes the variance in secondary cases based on a Negative Binomial distribution, where smaller values indicate more heterogeneity and skew and large values approach a Poisson distribution. Estimated *k* values for SARS-CoV-2 remain uncertain, but are thought to range from 0.04 − 0.3^6,7,11,12,^ similar to the estimate of 0.16 for SARS-CoV-1^2^, which we use for this analysis. These empirical and branching process approaches are ideal for characterizing heterogeneity in secondary cases, but not for projecting epidemic trajectories through time, without being further embedded in a compartmental or network modeling framework.

Here, we present a mechanistic susceptible, exposed, infectious, removed (SEIR) model that uses data on cases, deaths, and mobility for parameter estimation, incorporates heterogeneity in transmission rates, and is realistic enough to be useful for scenario exploration but simple enough to be adapted to a wide range of settings. The key innovation in our model is in using the average of Gamma-distributed *individual* transmission rates at each time step, as supported by previous work on secondary case distributions, to generate the distribution of *population-average* transmission rates. This formulation allows us to both generate more realistic variation in trajectories than SEIR models that assume a single average transmission rate, and explore and quantify the impact of altering individual-level transmission distributions on population-level dynamics without more detailed information on contact networks, age structure, or other social information.

The model, with accompanying open-access code, can be used to fit to any county in the U.S. using publicly available data; here we focus on five contrasting epidemiological settings—Seattle (King County), Washington; Los Angeles (Los Angeles County), California; Santa Clara County, California; Atlanta (Dekalb and Fulton Counties), Georgia; and Miami (Miami-Dade County), Florida. For each location we estimate a time-varying effective reproduction number, ℛ_*E*_, which represents the average number of secondary infections produced by each infected person, and is an important (though imperfect ^7^) metric of epidemic control. Using each fitted model, we truncate the individual-level transmission rate distribution and stochastically simulate epidemic dynamics into the future, representing a scenario where high-risk events are eliminated but smaller and lower-risk activities are allowed to resume. We investigate the absolute impact of this superspreading prevention strategy on epidemic control, and compare its impact on epidemic dynamics (and ℛ_*E*_) to test-and-isolate and shelter-in-place interventions. Using this comparison we highlight exit strategies from shelter-in-place that are expected to reduce both epidemic growth (i.e., keep ℛ_*E*_ below one) and the probability of explosive resurgence.

## Methods

### Model Structure

We developed a compartmental model using an SEIR (Susceptible, Exposed, Infectious, Recovered) framework to model COVID-19 transmission, which was first described in Childs et al. ^13^. Our model divides the population into the following classes: susceptible (S); exposed but not yet infectious (E); infectious and presymptomatic (I_P_), asymptomatic (I_A_), mildly symptomatic (I_M_), or severely symptomatic (I_S_); hospitalized cases that will recover (H_R_) or die (H_D_); recovered and immune (R); and dead (D). We assume an underlying, unobserved process model of SARS-CoV-2 transmission described by equations 1–10 and shown in Figure S1, where each term *d*_*X,Y*_ denotes the transition from compartment *X* to *Y*. Transitions between compartments are simulated as binomial (ℬ) or multinomial (M) processes. We use an Euler approximation of the continuous time process with a time step of 4 hours. To produce more realistic latent and infectious periods we divide each infectious class and the exposed period into multiple sub-stages, which results in Erlang distributed periods within stages ^14,15^. Specifically, we use three sub-stages for the exposed class, seven sub-stages for the asymptomatic infectious class, two sub-stages for the presymptomatic infectious class, five sub-stages for the mildly symptomatic infectious class, and five sub-stages for the severely symptomatic class. We translate durations into rates for our model with sub-classes and a Euler approximation using the method described in He et al. ^16^. Equations 11–18 describe in detail the stochastic rates used to approximate the transition terms in equations 1–10. Parameters are defined in Tables 1, 2, and 3.

**Table 1:** Parameter point estimates.

| *Parameter* | *Value* | *Description* | *Estimates and Sources* |
| --- | --- | --- | --- |
| $\kappa_P, \kappa_M, \kappa_S$ | 1 | Relative infectiousness of presymptomatic, mild symptomatic, and severe symptomatic | Assumed |
| $\gamma$ | 3.5 days | Preinfectious period | One meta-analysis<sup>19</sup> found a mean incubation period of 5.8 days, another found a median of 5.1 days<sup>20</sup>. We use a shortened duration because we assume 2 days of presymptomatic transmission |
| $\lambda_P$ | 2 days | Presymptomatic duration | Range of 1-3 days<sup>21</sup>, mean of 3.8 days<sup>22</sup>, viral shedding estimated to begin 2.3 days prior to symptom onset<sup>23</sup>, many articles find presymptomatic infection is likely but do not estimate duration<sup>24,25,26,27</sup> |
| $\lambda_A$ | 7 days | Infectious period for asymptomatic infections | Mean seroconversion after 7 days<sup>28</sup> |
| $\lambda_S$ | 5 days | Time from symptom onset to hospitalizations (severe cases) | Mean of 5.5 days<sup>29</sup>, median of 4 days<sup>30</sup>, mean of 5.6 days<sup>31</sup> |
| $\lambda_M$ | 5.0 days | Time from symptom onset to recovery (mild cases) | Infectiousness based on viral shedding estimated to decline substantially within 7 days<sup>23,28</sup> *Note,<sup>23</sup> takes samples from hospitalized patients; we assume similar viral shedding in mild infections |
| $\rho_R$ | 13.3 days | Time from hospitalization to recovery | Mean of 13.3<sup>32</sup>, mean of 20.51<sup>31</sup>, highly variable by region<sup>33</sup> |

**Table 2:** Parameter range estimates that are not location specific

| *Parameter* | *Lower Bound* | *Upper Bound* | *Description* | *Estimates and Sources* |
| --- | --- | --- | --- | --- |
| $\kappa_S$ | 0.4 | 0.8 | Relative infectiousness of asymptomatic infections | 0.6<sup>34</sup>, few direct estimates, but many examples of asymptomatic transmission potential less than, but potentially close to that of, symptomatic infected individuals<sup>35,36,37</sup> |
| $\alpha$ | 0.3 | 0.5 | Proportion of infections that are asymptomatic | Mean of 43.3%<sup>38</sup>, 44%<sup>39</sup> |
| $\delta$ | 0.1 | 0.3 | Fatality rate among hospitalizations | Demographic weighted average that will vary by location, see Verity et al.<sup>31</sup>, 5%<sup>40</sup> |
| $\rho_D$ | 13 days | 20 days | Time from hospitalization to death | Mean of 16 days<sup>41</sup>, mean of 17.8 days<sup>31</sup>, highly variable by region<sup>33</sup> |
| $1 - \mu$ | 0.025 | 0.075 | Proportion of symptomatic infections that require hospitalization | Demographic weighted average that will vary by location, see Verity et al.<sup>31</sup> |

**Table 3:** Location-specific parameter range estimates. Population sizes obtained from the US Census Bereau ^42^

| Parameter | Santa Clara County, CA | Los Angeles County, CA | Miami-Dade County, FL | King County, WA | Fulton+DeKalb, GA |
| --- | --- | --- | --- | --- | --- |
| Epidemic Start Date | 01-Jan - 05-Feb | 01-Jan - 31-Jan | 01-Jan - 16-Mar | 01-Jan - 04-Mar | 01-Jan - Mar-07 |
| Population Size | 1,927,852 | 10,039,107 | 2,716,940 | 2,252,782 | 1,755,830 |

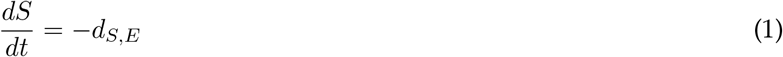

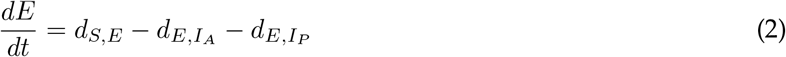

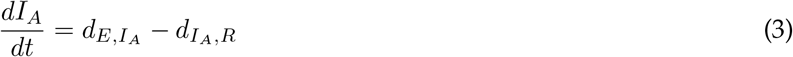

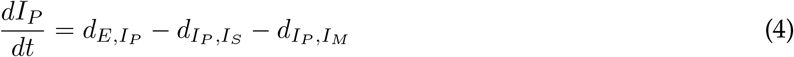

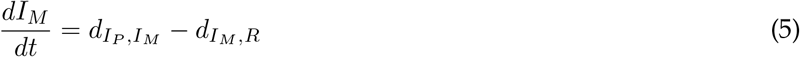

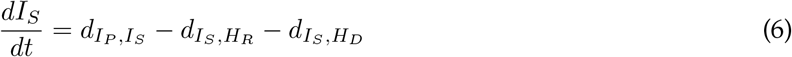

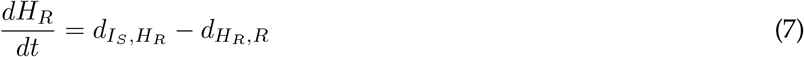

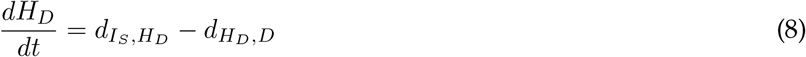

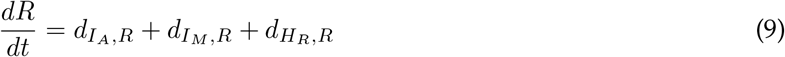

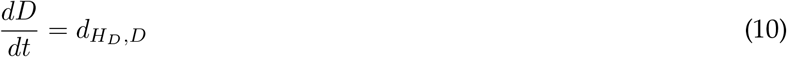

By including asymptomatic and presymptomatic individuals, we are able to track “silent spreaders” of the disease, which have been shown to contribute to COVID-19 transmission ^17,18^. Mildly symptomatic cases are defined as those people that show symptoms but do not require hospitalization. We assume that all severely symptomatic cases will eventually require hospitalization and that no onward transmission occurs from hospitalized individuals.

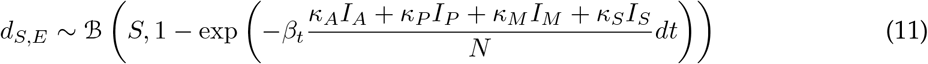

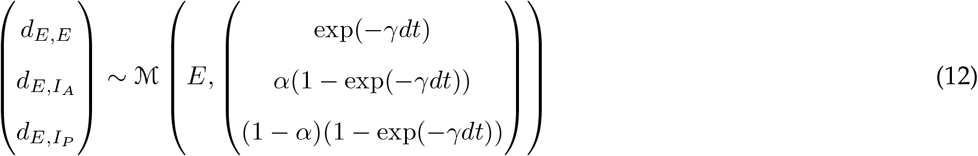

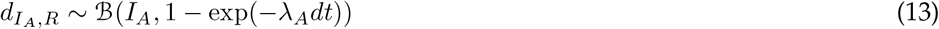

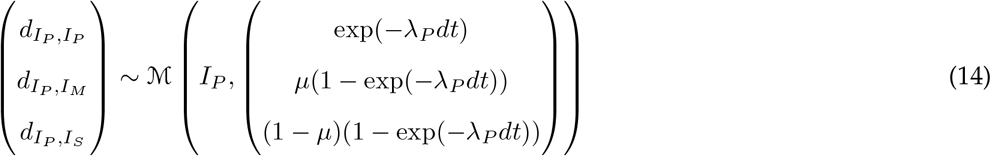

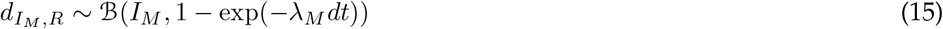

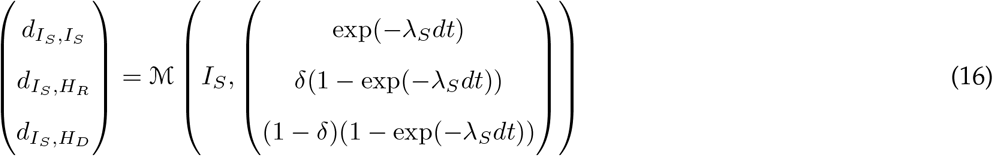

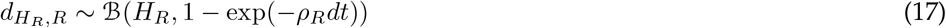

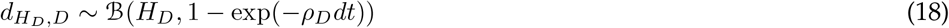

The time-varying transmission parameter, *β*_*t*_, describes the average per capita rate of contact between susceptible and infectious people at time *t*, multiplied by the per-contact transmission probability. We modeled *β*_*t*_ as a function of human movement using the scaling function:

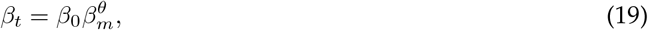

which treats *β*_*t*_ as an exponentially decreasing function of physical distancing (*θ*; on a scale of 0-1 where 0 is no physical distancing, and 1 is maximum physical distancing). Here, *β*_0_*β*_*m*_ is the estimated minimum possible transmission rate given minimal human movement (i.e., maximal physical distancing) and thus contact rate. To model human movement we use SafeGraph’s “Shelter in Place Index” ^43^, which measures the proportion of cell phone devices that are staying home.

To model individual heterogeneity in SARS-CoV-2 transmission rate, we allow individuals to vary over time by modeling an individual’s transmission rate in each time step as a Gamma distributed random variable with a dispersion such that the sum of an individual’s transmission rates over the duration of their infection approximates a Gamma distributed random variable with dispersion equal to previous Negative Binomial parameterizations for reproductive number SARS-CoV-1 (*k* = 0.16) ^44^, which closely approximates estimates of overdispersion for SARS-CoV-2^12,45^. Because we model the transmission rates as the multiplication of contact rate and infection probability, this heterogeneity implicitly considers both variation among individuals in infectiousness and contact rate, and can be thought of as modeling superspreading *periods or events*—windows in time when an infected individual has a particularly high transmission rate. To incorporate this variation into an average time step *β*_*t*_, we model *β*_*t*_ as the average of the transmission of all infected individuals at time *t*. To do so we apply the property of Gamma distributions that the mean and variance of *N* samples from a Gamma distribution with defined rate and scale is itself a Gamma distribution with mean equal to that of the original Gamma distribution and variance equal to the variance of the original Gamma divided by *N*. A full derivation of the equivalence between the individual time step transmission rate distributions (which we will hereafter refer to with *π*), the individual infectious period transmission rate distributions, and the population-level transmission rate distribution is available in the Appendix.

We assume that observed deaths are a Negative Binomial random variable with a mean equal to total new deaths accumulated over the observation period (i.e., one day for this analysis), and a dispersion parameter that we estimate. We also assume that daily observed cases are a Negative Binomial random variable, but have a mean equal to the daily number of new symptomatic infections multiplied by a daily detection probability that we estimate from the data. We model daily detection probability as a monotonically increasing logistic function:

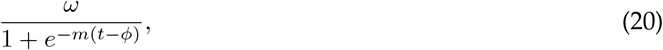

where *ω* is the maximum fraction of symptomatic cases detected, *m* is the logistic growth rate, and *φ* gives the location of the inflection point (where the probability of detection equals one half of the maximum detection probability, *ω*). Because *φ* can be estimated to be in the future, the probability of detection of an infected case in the present can be any value between 0 and *ω*. We estimate newly observed cases to be a fraction of all new symptomatic infections at time *t*. Though this ignores testing asymptomatic infections, any detection of asymptomatic infections will be captured as a higher estimated detection fraction of symptomatic infections.

### Fitting the Model

We use COVID-19 death and case data from The New York Times, based on reports from state and local health agencies (available at https://github.com/nytimes/covid-19-data). Using these data, which are available for all counties in the US, and any form of human movement data that can be scaled to 0-1, our model can be used to fit infection dynamics in any county.

For computational efficiency, we assumed point estimates for some parameters (Table 1) and sampled over uncertainty in others (Tables 2, 3) by drawing 600 sobol sequences, an efficient method for sampling input parameters ^46^, across a range of plausible values for each in order to form 600 plausible parameter sets. For each parameter set we used the package `pomp` ^47^ in the statistical programming language `R` ^48^ to estimate the following parameters: *β*_0_: transmission rate over an entire infection in the absence of social distancing; *β*_*m*_: estimated transmission given zero human movement; *E*_0_: number of exposed individuals that initiate the epidemic; *ω, m*, and *φ*: maximum, slope, and inflection point day of the sigmoidal case detection function; *θ*_*d*_: Negative Binomial dispersion parameter for deaths; and *θ*_*c*_: Negative Binomial dispersion parameter for cases. We fit all parameters to daily deaths, cases, and mobility in two steps. First, for each of the 600 parameter sets we used the `mif2` function in `pomp` with random starting conditions, 120 iterations and 2000 particles. We then continued to fit the 60 parameter sets with the highest log likelihoods for an additional 200 iterations using 2000 particles. Each county took approximately nine hours to fit using twenty cores.

We calculated ℛ_*E*_ at each time *t* as estimated *β*_*t*_ times the median proportion of the population remaining susceptible on each day across 300 simulated epidemics, with simulated epidemics that did not reach at least a total of 100 infected discarded, times the average infectiousness over an infection (as defined by our model structure) using the 10 parameter sets with the largest negative log likelihoods as determined by the second fitting step.

### Simulating epidemics under interventions

Any intervention type, intensity, or duration can be modeled using this framework and open-source code (available at https://github.com/morgankain/COVID_interventions) given that it can be written as a function that modifies either human movement or *β*_*t*_ (e.g., social distancing or a pharmaceutical intervention that reduces the probability of infection). Previously we considered the impacts of various social distancing initiatives on epidemic dynamics using a similar model formulation ^13^. Here we consider interventions that reduce the skew of the individual time step transmission rate distribution (*π*), and thus the average time-varying transmission rate *β*_*t*_; this is our mathematical representation of reducing highly infectious contact periods or events, which for COVID-19 tend to occur in crowded enclosed environments (e.g., church choirs and exercise classes). Specifically, we model truncation of the *π* distribution by assuming that all samples within the top X% of the *π* distribution are resampled. To visualize the dynamics of interventions, for each location we simulate 300 epidemics from the maximum likelihood estimate across the 600 parameter sets. The uncertainty band we plot represents the central 95% range of outcomes seen across all stochastic realizations that resulted in epidemics for this parameter set, and thus should not be taken as representation of uncertainty in parameter values or model structure.

## Results

### Epidemic trajectories

The model produced realistic fits to five contrasting epidemiological settings—King County, Washington; Los Angeles County, California; Santa Clara County, California; Fulton and Dekalb Counties, Georgia; and Miami-Dade County, Florida (hereafter, Seattle, Los Angeles, Santa Clara County, Atlanta, and Miami). Among these locations, we estimated that prior to interventions, ℛ_0_ ranged between approximately 2 and 4 (Figure 1). We also estimated that ℛ_*E*_ dropped below one following shelter-in-place orders in all counties, though only briefly in some locations. In particular, in Miami, Los Angeles, and Atlanta ℛ_*E*_ climbed above 1 by mid-May and daily cases and deaths have plateaued or continue to grow into June. Though ℛ_*E*_ remained below 1 into at least early June in Seattle and Santa Clara County, as of June 18 ℛ_*E*_ is *∼*1 and cases are rising again.

**Figure 1:**
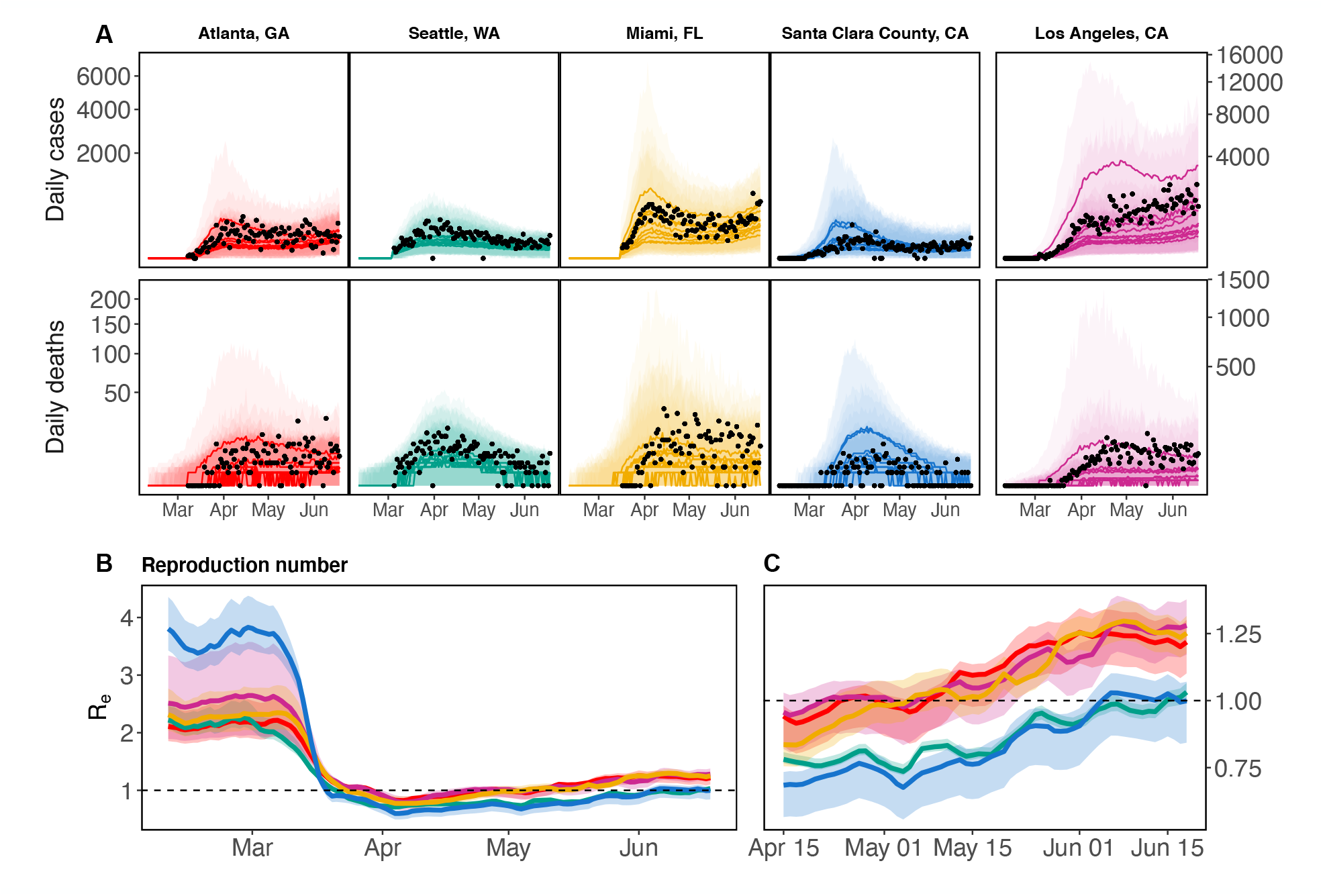
Model estimated daily cases and deaths (A), and reproduction number (B, C) for five locations: Atlanta (red), Seattle (green), Miami (gold), Santa Clara County (blue), and Los Angeles (purple). Los Angeles is displayed on a different *y*-axis due to differences in magnitude of reported deaths and cases. For each county, we show the 10 model fits with the best log likelihoods. Panel C show the same results pictured in B, but are zoomed in to April 15 - June 18 to better show the dynamics around ℛ_*E*_ = 1. Black points are observed daily deaths and reported cases in each county. Solid lines display mean of model simulated trajectories (A) and mean *R*_*e*_ (B, C). Ribbons show the range of estimated *R*_*e*_ (B, C) or 95% CIs over stochastic simulation from each model fit (A). Vertical axes in panel A are square root transformed for visibility.

### Interventions

As a basis for comparison, focusing on just two locations—Los Angeles and Seattle—if shelter-in-place were simply lifted, a second peak would be inevitable in the absence of any non-pharmaceutical interventions (Figure 2, blue). However, non-pharmaceutical interventions, including continuing shelter-in-place, infected isolation with intermediate levels of shelter-in-place, or averting superspreading with intermediate levels of shelter-in-place are capable of limiting epidemic growth (Figure 2) and keeping ℛ_*E*_ near or under 1. Here, we consider intermediate levels of shelter-in-place that correspond to mobility levels that are an average of baseline mobility prior to social distancing and final mobility levels observed in the last week of data. Either an infected isolation strategy that reduces to intermediate levels of shelter-in-place and catches 90% of all mild and severe cases of COVID-19 before they are able to transmit (Figure 2, green), or a truncation strategy that similarly reduces to intermediate levels of shelter-in-place but removes the top 1% of the individual time step transmission rate distribution (*π*) with 75% efficiency (Figure 2, purple) are able to suppress epidemic growth (and reduce ℛ_*E*_ to below one) in Los Angeles, CA and Seattle, WA.

**Figure 2:**
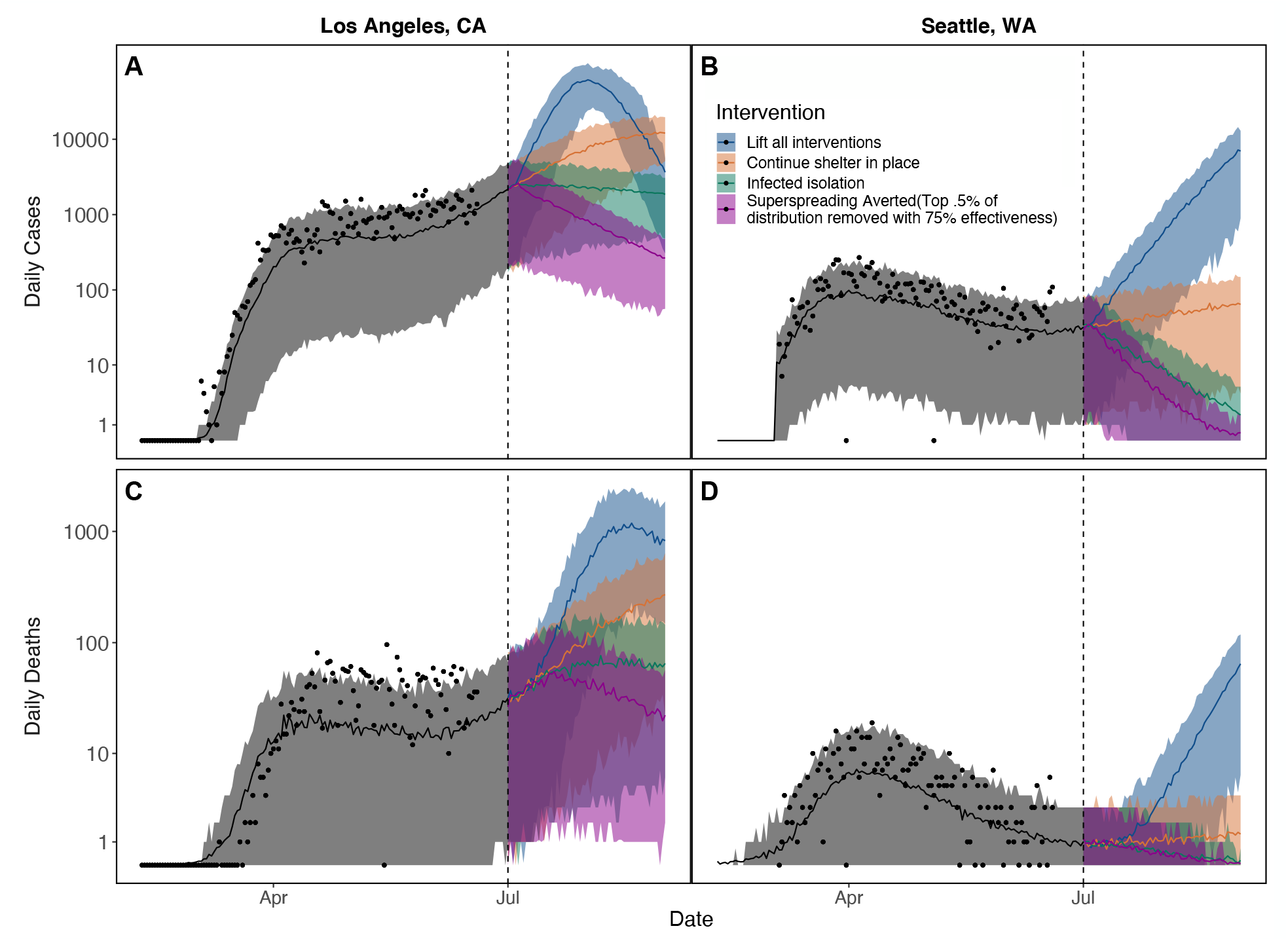
Maintaining shelter-in-place (SIP; orange), test-and-isolate (green), or superspreading aversion (purple) strategies over long periods is necessary to prevent a major epidemic resurgence (blue) in each location where we fit our model (shown here for Los Angeles, CA [A, C] and Seattle, WA [B, D]). However, continuing SIP at current levels (orange) will lead to an increase in daily cases in both Los Angeles (A) and Seattle (B). Daily reported cases are shown in (A) and (B) and daily deaths in (C) and (D). For both shelter-in-place and truncation interventions we assume an intermediate level of mobility (an average of baseline mobility prior to social distancing and final mobility levels observed in the last week of data). Bands show 95% CI on stochastic simulations of daily cases and deaths for the single maximum likelihood parameter set. Dates range from February through September of 2020. Vertical axes are log transformed for visibility.

### Curtailing superspreading

Limiting opportunities for superspreading by “chopping off the tail” of the contact rate or infectiousness distributions can be highly effective at epidemic control (Figure 2), driving epidemic growth to be negative and bringing the average number of secondary cases (ℛ_*E*_) below 1. An example truncation intervention is illustrated in Figure 3: because the individual transmission rate distribution, *π*, over a 4-hour time period is so skewed (Figure 3A; see appendix for derivation), truncating the upper 0.1% yields a large reduction in the mean and a moderate reduction in the variance of the population-level average transmission rate (Figure 3B, shifting from red to blue distribution). A variety of possible truncation strategies exist, including eliminating varying proportions of *π* (e.g., upper 0.5%, upper 1%) with varying levels of efficiency (ranging from 50-100%) (Figure S2).

**Figure 3:**
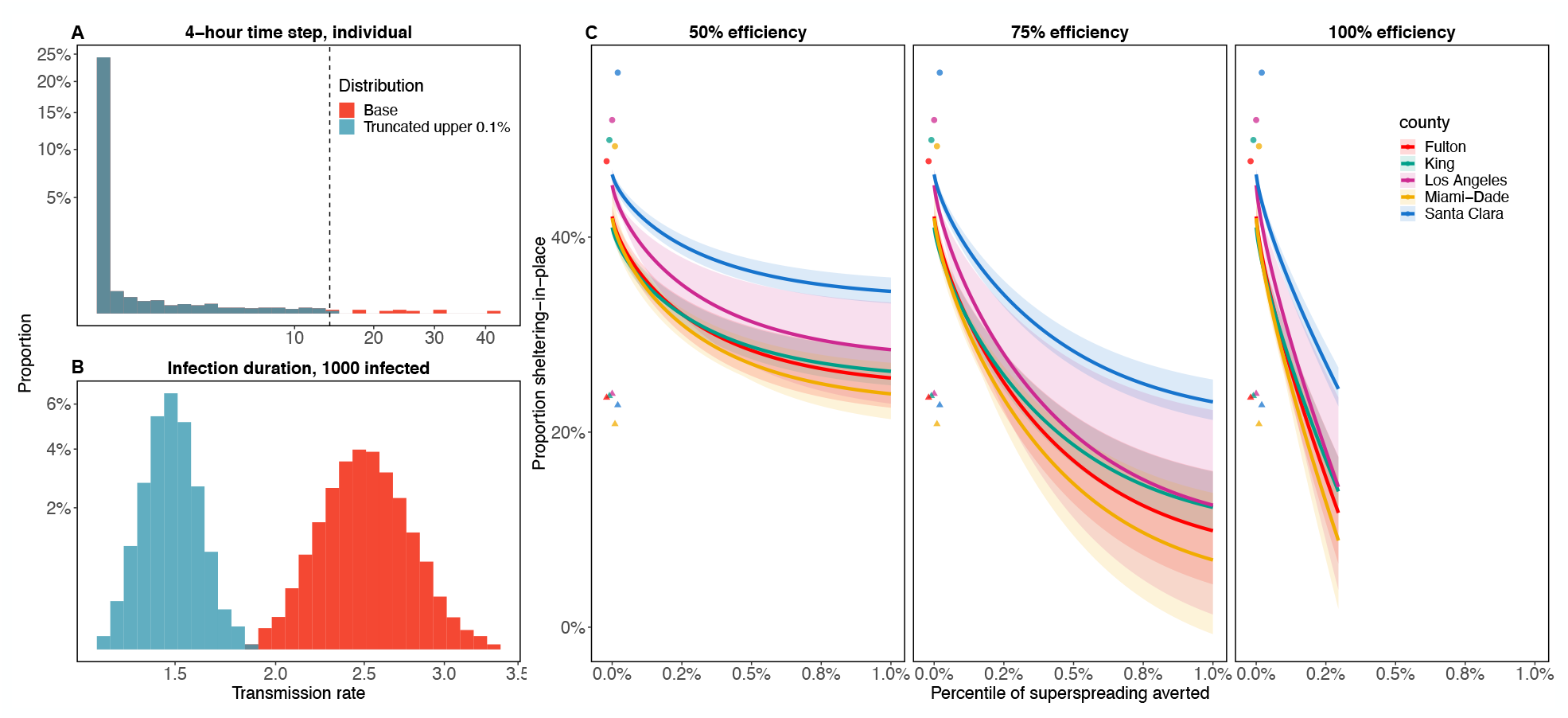
Example of how truncating the individual-level transmission rate distribution, *π*, (A) affects the population-average transmission rate (B), and combinations of sheltering-in-place (SIP) and truncation strategies that reduce ℛ_*E*_ to one in a fully susceptible population (C). The three panels in C show the combinations of truncation and SIP that produce an ℛ_*E*_ of one for three levels of truncation efficiency. (A) Truncation at the upper 0.1% of *π* (sampled over a 4-hour time step), in which truncation occurs for all values above the dashed line. (B) Resulting effect on the population-level average infection rate when there are 1000 infected people currently in the population, where the original distribution is in red and the truncated distribution is in blue. The distribution is shown over 10,000 simulations for a population characterized by an individual reproduction number distribution with mean of 2.5 and overdispersion parameter, *k* = 0.16. Horizontal and vertical axes in A and B are square root transformed for visibility. In C, the triangles show baseline SIP in each location and circles show max SIP reached during social distancing. Solid lines indicate the mean over the ten best fits, and the ribbon is the full range of estimates from these fits.

An alternative measure of the impact of averting superspreading (i.e., truncation interventions) is how much general social distancing can be avoided by instead truncating the transmission rate distribution. Prior to social distancing orders, the estimated proportion sheltering in place (SIP, for short) ranged from *∼*20-22% across our focal locations (Figure 3C, triangles), indicating the baseline level of mobility. If we combine SIP with truncation interventions, a variety of combinations are predicted to provide epidemic control (for example, by reducing transmission rates such that ℛ_*E*_ in a fully susceptible population would be 1; Figure 3). If the truncation intervention is 100% effective, truncating only approximately the upper 0.15% of individual transmission rates, *π*, (Figure 3A) is effective enough to maintain transmission rates such that ℛ_*E*_ would be 1 in a fully susceptible population, while allowing mobility levels to return to baseline (Figure 3C). Alternatively, if truncation interventions are only half as effective, the same 0.15% truncation intervention would require moderate-strong social distancing (SIP from *∼*30-45%; Figure 3C). The nonlinear effects of social distancing and truncation on transmission make the combination of interventions needed to maintain epidemic control sensitive to the efficiency and strength of each intervention mechanism.

### Superspreading and epidemic resurgence

Even if the epidemic is brought almost entirely under control (e.g., to within 1-5 infected individuals remaining in the population), epidemic resurgence remains a possibility if interventions wane, allowing ℛ_*E*_ to increase above one. As we show in (Figure 3), many different combinations of SIP and truncation can be used to produce the same ℛ_*E*_ (in Figure 3, an ℛ_*E*_ of 1); however, epidemic dynamics will vary by combination because of the variation in individual time-step transmission rates, *π*. If ℛ_*E*_ rises above one because interventions are relaxed, the specific combination of SIP and truncation that remains in place will determine the resulting dynamics. Here we examine how different truncation interventions will affect epidemic extinction probability and the size of epidemic resurgence when it does not go extinct. We compare the full effect of truncation interventions (which influence both the mean of the transmission rate distribution and its shape) to the effects of truncation when ℛ_*E*_ is held constant by scaling SIP, reflecting only truncation effects on the transmission rate distribution shape (variance, skew, etc.).

With few infected individuals and ℛ_*E*_ *>* 1, stochasticity and heterogeneity in *β*_*t*_ can either lead to extinction, moderate resurgence, or explosive resurgence. Keeping interventions in place that remove even a tiny percent of the largest *β*_*t*_ values can help to avoid the more explosive events (Figure 4). Truncation markedly reduces the probability of explosive epidemic resurgence (Figure 4A) both by increasing the extinction probability (Figure 4B) and by reducing the magnitude of resurgent epidemics when they do occur (Figure 4C). While epidemic size was less sensitive to the number initially infected when resurgences do occur (Figure 4C), the stochastic extinction probability was extremely sensitive to the difference between even one, three, or five remaining infections (Figure 4B). Much, but not all, of the benefit of truncation comes from changing the mean transmission rate (and therefore ℛ_*E*_). When ℛ_*E*_ is held constant by adjusting SIP, effects of truncation are more moderate. An increase in efficiency at truncating the top 0.1% of the *β*_*t*_ distribution noticeably decreases the number of infected 42 days after interventions are relaxed (Figure 4D,F). However, because of the need to slightly reduce SIP to hold ℛ_*E*_ constant under truncation, truncation of *π* marginally decreases the extinction probability of the epidemic, which remains much more sensitive to the number initially infected (Figure 4E). The highly stochastic nature of epidemic growth when cases are rare, combined with the fact that each truncation leaves behind highly skewed distributions regardless of the truncation parameters, results in even 10,000 epidemic simulations producing noisy patterns across intervention scenarios. Similar patterns are seen as more of the *π* distribution is truncated (Figure S4).

**Figure 4:**
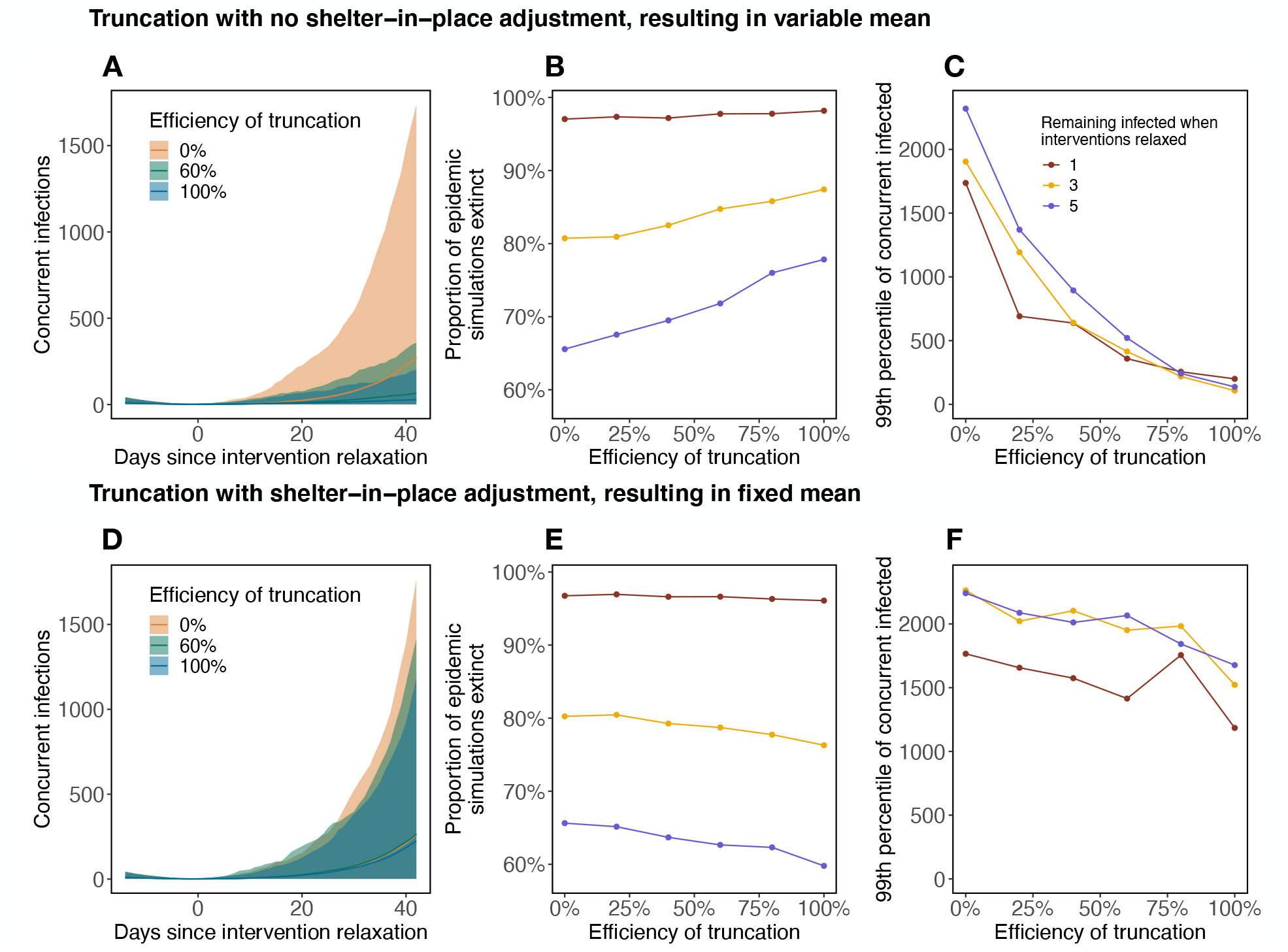
Effects of transmission rate truncation on epidemic die-out and explosive resurgence. With skewed individual variation in transmission rate, relaxing social distancing interventions when infections become rare (allowing ℛ_*E*_ to increase above one) may lead to explosive stochastic epidemic resurgence. Top panels (A-C) show the overall effect of truncation interventions, including effects on both the mean and shape of the transmission rate distribution, and resulting ℛ_*E*_. Bottom panels (D-F) show the effect of truncation when ℛ_*E*_ is held constant by rescaling shelter-in-place at the time of intervention relaxation. Specifically, for a 0% truncation efficiency we simulate epidemic resurgence assuming ℛ_0_ = 2, which results in an ℛ_*E*_ = 2 *S/N* at the time of resurgence, which will vary by simulation (where *S* is the number of susceptible individuals and *N* is the total population size). In panels (A-C) as truncation efficiency increases ℛ_*E*_ decreases; in panels (D-E) we scale shelter-in-place to retain an average ℛ_*E*_ = 2 *S/N* across truncations. Simulations are performed with varying efficiencies of truncation of the top 0.1% of the *π* distribution. Envelopes in (A) and (D) show the central 98% of resurgent simulations (across 10,000 total simulations) for three efficiencies of truncation (0% in orange, 60% in green, 100% in blue). The proportion of epidemic simulations that go extinct within 42 days of intervention relaxation for thresholds of 1 (red), 3 (gold), and 5 (blue) infected individuals is shown in (B) and (E). The upper 99th percentile of concurrent infections 42 days after intervention relaxation in resurgent simulations for the same thresholds is shown in (C) and (F).

## Discussion

Understanding local epidemiological dynamics of COVID-19—and the impact of heterogeneity on those dynamics—remains a challenge due to both limited and imperfect data in most regions and ever evolving interventions and adherence. Reported cases are only a small fraction of all infections, and the proportion of symptomatic cases that are detected remains highly uncertain and variable over space and time. Our approach takes an important step toward capturing locally-specific epidemic dynamics and the impact of heterogeneity across settings by providing a platform (including a mathematical model and open access code) for estimating time-varying transmission rates (*β*_*t*_) from death, mobility, and imperfectly observed case report data, all of which are publicly available. The model can estimate epidemic dynamics and transmission rates over time across epidemiological settings that vary in population size, demography, and control. By incorporating individual variation in contact rates (or, equivalently, infectiousness) into time step transmission rate distributions, we incorporate some of the known effects of heterogeneity without requiring detailed information on population mixing, structure, social networks, or movement patterns. We find that control measures in March of 2020 rapidly brought the average reproduction number—ℛ_*E*_—from *∼*2–4 to below 1 in all locations we considered in early April. However, as of June 18, ℛ_*E*_ has once again drifted above one in all of these locations except possibly in Seattle, WA and Santa Clara County, CA, where it remains unclear if it is greater or less than one.

Non-pharmaceutical interventions will be necessary to control COVID-19 in all settings until better pharmaceutical options (in particular, effective vaccines) are widely available. Social distancing in the general population is effective but costly: it is a blunt and imprecise tool. The social and economic necessity of relaxing social distancing demands safe exit strategies based on more precise, targeted interventions to reduce transmission. Testing and isolating symptomatic people, combined with contact tracing, remains the gold standard intervention for limiting onward transmission as social distancing is lifted, but it is expensive and capacity remains limited in many settings. Our model shows that it is possible to target interventions even without precise information on specific population mobility, mixing, and infectiousness patterns, by limiting just the most high-risk activities, such as large gatherings and indoor events that have many close contacts. How much can be gained from these common sense interventions that reduce or eliminate opportunities for superspreading while allowing smaller and safer activities to resume? We find that these truncation interventions, which eliminate the upper percentiles of contact rates in the population, and thereby transmission rates, can be highly effective at maintaining epidemic control (Figure 2), particularly when combined with mild to moderate social distancing (Figure 3). Importantly, even after epidemic control is achieved and case numbers drop very low, “chopping off the tail” can provide powerful insurance against explosive resurgence after social distancing interventions are otherwise lifted (Figure 4).

What does “chopping off the tail” mean in practical terms? Five types of factors tend to promote superspreading: (1) high rates or intensity of contact between people or with surfaces; (2) large aggregations of people; (3) poorly ventilated physical environments, especially indoors ^49^; (4) highly infectious individuals; (5) highly susceptible recipient population ^4,6,50^. Many settings where SARS-CoV-2 superspreading has occurred—including nursing homes ^26^, exercise classes, bars and restaurants ^49^, funerals, churches ^51^, meat-packing plants ^52^—combine multiple risk factors. For example, choir practices combine high densities of people, a high-risk activity (singing) ^53^, and potentially poorly ventilated indoor spaces; long-term care facilities combine mobile, high-contact caregivers with highly vulnerable residents, often in high-density indoor spaces. Some superspreading events may be easier to eliminate than others. Clearly, healthcare and long-term care facilities serve critical functions despite their high-risk nature, and taking all possible steps for decontamination and personal protection in these facilities is critical to mitigate this risk ^50^. On the other end of the spectrum, voluntary, large, indoor events that are mainly for entertainment and could be postponed—gyms, clubs, sporting events, concerts, large lectures—may be the most viable option to reduce superspreading and “chop off the tail” of the contact rate distribution ^54^. While these common sense interventions are not novel suggestions ^6^, and are already part of reopening plans in almost all locations, our work allows a direct comparison of how much general social distancing is avoided by eliminating a fraction of these high-risk events (Figure 3). Truncation strategies are even more desirable in light of their effectiveness at preventing explosive resurgence after controls are otherwise lifted (Figure 4). Mapping actual event types onto the contact rate distribution to determine how particular superspreading reduction policies would affect control remains an important next step. Importantly, associating superspreading with events and locations, rather than specific people, can avoid the stigma sometimes associated with being identified as a superspreader ^4^.

The impact of truncation interventions is two-fold. First, removing the upper tail of the individual transmission rate distribution reduces the population-level mean, often dramatically (Figure 3A,B). If the mean transmission rate already placed ℛ_*E*_ near 1 (for example, due to other interventions), then additional truncation could be enough to cross this critical threshold. However, most intervention strategies that bring ℛ_*E*_ to 1 already include prohibiting large gatherings, especially indoors, so additional truncation may not be possible within the context of first-wave interventions. However, truncation also acts on the variance and skew of the transmission rate distribution, though these effects are smaller than the effect on the mean (Figure 4D-F compared to A-C). Given that super-spreading events are particularly dangerous when cases are few (in the early or late phases of the epidemic) ^2^, sustained truncation interventions could be extremely important for preventing explosive stochastic re-emergence when low case numbers allow general social distancing to be lifted (Figure 4). In this scenario, resurgence remains rare (Figure 4B) but possible because individual variation in transmission rates is large; most of the time infectious people transmit to few others, but occasionally someone infects dozens (Figure 4A), quickly overwhelming testing, contact tracing, and isolation efforts. Sustained truncation dramatically reduces the probability of explosive resurgence, and constrains incipient transmission chains to be smaller and more manageable.

One limitation on understanding the effect of heterogeneity in transmission in particular locations is the challenge of estimating epidemiological parameters from noisy and imperfect data: necessarily a balancing act between model simplicity and complexity. Here, we rely on metrics of heterogeneity previously estimated for SARS-CoV-1 and SARS-CoV-2^2,12,45^ instead of estimating them directly from data; we focus our parameter estimation on the mean of the transmission rate distribution. Heterogeneity in contact rates or infectiousness, and the resulting distributional variance and skew, may vary based on local patterns of movement, contact, behavior, and population demography. This heterogeneity can have important consequences: in some cases epidemics with low mean *R*_0_ can actually infect a larger proportion of the population than epidemics with higher mean *R*_0_—as was the case for the 1918 influenza pandemic as compared to the 2014 Ebola outbreak—due to the heterogeneity in transmission rates, as described by higher moments of the secondary case distribution ^7^. The true epidemiological parameters in any given location, and the extent of our uncertainty in these parameters, also remain unknown because of the computational challenges of parameter estimation given the limited information contained in noisy case, death, and mobility data. For example, depending on how a particular candidate parameter combination weights the noisiness of cases and deaths and estimates initial conditions, transmission rate estimates can vary substantially (Figure 1). Fully characterizing uncertainty in model structure and parameter values in this context is difficult. Future work that directly estimates case ascertainment rates (e.g., through metrics of percentage of tests that are positive, age distributions of positive tests, epidemiological contact information on cases, and analysis of viral genome sequences ^55^), as well as more detailed mobility and contact network information ^17^ could help to improve the model fit to the full shape of the transmission rate distribution.

First-wave interventions that eliminated large social gatherings and indoor activities and mandated mask-wearing and physical distancing have likely already affected the heterogeneity in transmission rates, by eliminating many of the high-risk events likely to fall into the upper tail of the distribution. It is important to recognize that as social distancing interventions relax, sustaining such truncation interventions may be critical for keeping transmission down to levels manageable through testing, contact tracing, and isolation. This truncation strategy can potentially reduce the social and economic costs of non-pharmaceutical interventions on the general populace, and facilitate sustained adherence by allowing lower-risk activities to resume while insuring against a resurgence. Ultimately, an unmitigated epidemic, whether as a first or second wave, would kill thousands to tens of thousands of people in each of the locations we studied, reinforcing the point that aiming for population herd immunity through naturally acquired infections is not a viable public health strategy. Instead, exit strategies that can sustain epidemic control after shelter-in-place orders end, including truncating the transmission rate distribution, will be necessary until an effective vaccine can be developed and widely distributed.

## Data Availability

All data used in this study is publicly available. Code is available on github.

https://github.com/morgankain/COVID_interventions

https://www.safegraph.com/dashboard/covid19-shelter-in-place?s=US&d=06-04-2020&t=counties&m=index

https://github.com/nytimes/covid-19-data

## Data and Code Availability

Data used in this study are available at: https://github.com/nytimes/covid-19-data. Code used to produce the results in this study are available at: https://github.com/morgankain/COVID_interventions.

## Acknowledgements

We thank the members of the Mordecai Lab at Stanford University for feedback on our model. Funding provided by: the National Science Foundation (DEB-1518681); the National Institutes of Health (National Institute of General Medical Sciences: R35GM133439); the Natural Capital Project; the Helman Scholarship; the Terman Award. Morgan Kain was supported by the Natural Capital Project. Marissa Childs was supported by the Illich-Sadowsky Fellowship through the Stanford Interdisciplinary Graduate Fellowship program at Stanford University.

## Author Contributions

MPK helped to design and code the model, conduct model fits and simulations, troubleshoot problems and update the model, write and revise the manuscript.

MLC helped to design and code the model, conduct model fits and simulations, troubleshoot problems and update the model, write and revise the manuscript.

ADB helped to refine the model, conduct model fits and simulations, troubleshoot problems and update the model, and revise the manuscript.

EAM helped to design the model, provide conceptual framing, troubleshoot problems, and write and revise the manuscript.

## Appendix

### Derivation of the population-level *β*_*t*_ **distribution**

In heterogeneous populations, the expected number of secondary infections caused by a particular individual (or the individual reproductive number, *ν*) can be modeled as a negative binomial random variable with mean *R*_0_ and overdispersion parameter *k* ^2,6,56^, i.e. 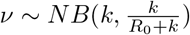. This is equivalent to modeling *ν* as a Poisson random variable whose mean is itself a random gamma variable with shape *k* and scale *R*_0_*/k*,

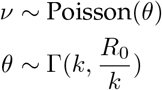

Now let *d* be the duration of infection for an individual and *τ* be a time step. Using the fact that *k* = Σ_*M*_ *k/M*, we have

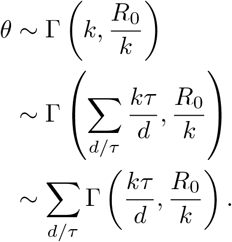

Thus for a constant duration of infection *d*, we have the individual infection rate over a time step

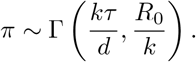

When there are *N* infected individuals, the average infection rate *β*_*t*_ over a time step is

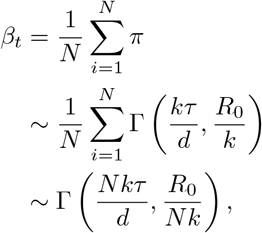

which will have mean *R_0_τ/d* and variance 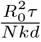. Notably, this behaves well with scalings on *R*_*0*_ as a function of interventions: Let *θ* be the amount of physical distancing occurring in the population on a scale of 0-1 where 0 is no physical distancing, and 1 is maximum physical distancing, and *f* be a function mapping *θ* to a scaling on *R*_0_. Now 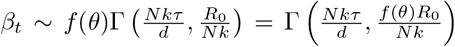, which means that properties of the distribution are preserved with *f* (*θ*)*R*_0_. Specified as a gamma white noise process Γ_*W N*_ (*σ, µ*) which has mean *µ* and variance *σ*^2^*µ*, this is equivalently

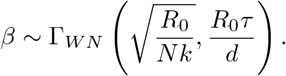

There are two main differences between the above derivation and our model formulation (note that for the following we assume *f* (*θ*) = 1):

1. above, the number of new infections in a time step should be

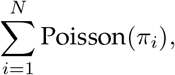

where each *π*_*i*_ is i.i.d. as 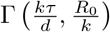. In our model the number of new infections in a time step is

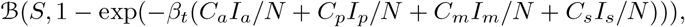

where *β*_*t*_ is the average transmission rate over all individuals infectious during that time step and ℬ is a binomial process. For large *S* and small *I*_*a*_ +*I*_*p*_ +*I*_*m*_ +*I*_*s*_, this approximates a Poisson distribution for the number of secondary cases from each infected individual in each time step and the total secondary infections caused by an individual over their infectious period.

2. above, we assume a constant duration of infection. In our model periods are Erlang distributed given our division of stages (e.g. *I*_*a*_) into *n* sub stages, each with the same period ^14,15^. This marginally increases the variance in the per-infectious period d istribution as we show in Figure S5.

### Derivation of the relationship between *R*, **Gamma truncation, and the fraction of individuals sheltering in place**

Let *d* be the average duration of infection, *τ* be a time step, *θ* be the proportion of the population sheltering in place, and *X*_*p,η*_ be a random variable *X* with right truncation, where truncation occurs at the *p*-th percentile, with probability *η*. Suppose that in a given time step we truncate the individual infection rate (*π*) over a time step at the *p*-th percentile with probability *η*. Then the reproduction number is

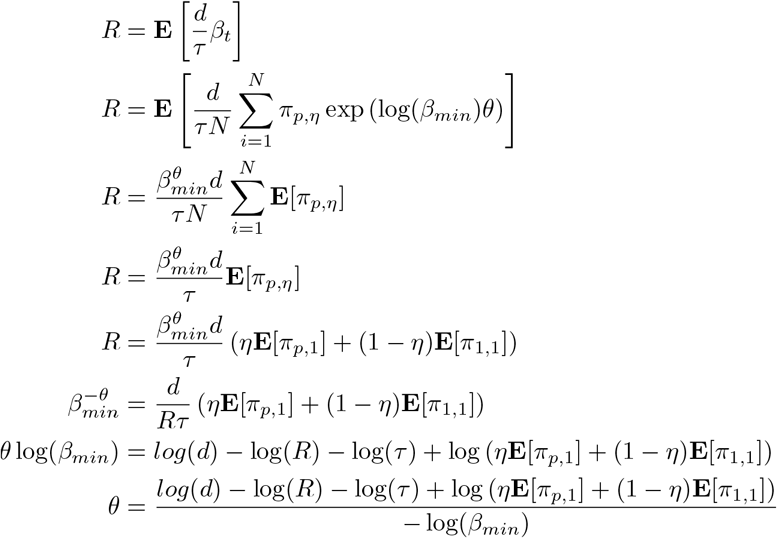

For a truncated gamma distribution with shape *a* and scale *b* with upper truncation at *u*, the expected value is

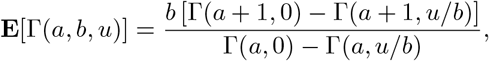

where Γ is the upper incomplete gamma function. See Okasha and Alqanoo (2014) [eq.29] ^57^ for the full derivation.

Letting *γ* be the lower incomplete gamma function, it follows that

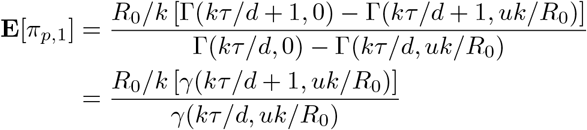

where *u* is the *p*-th percentile of *π* ^57^. *Then*

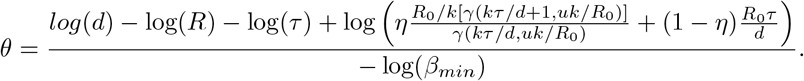

## Supplemental Material for “Chopping the tail: how preventing super-spreading can help to maintain COVID-19 control”

**Figure S1:**
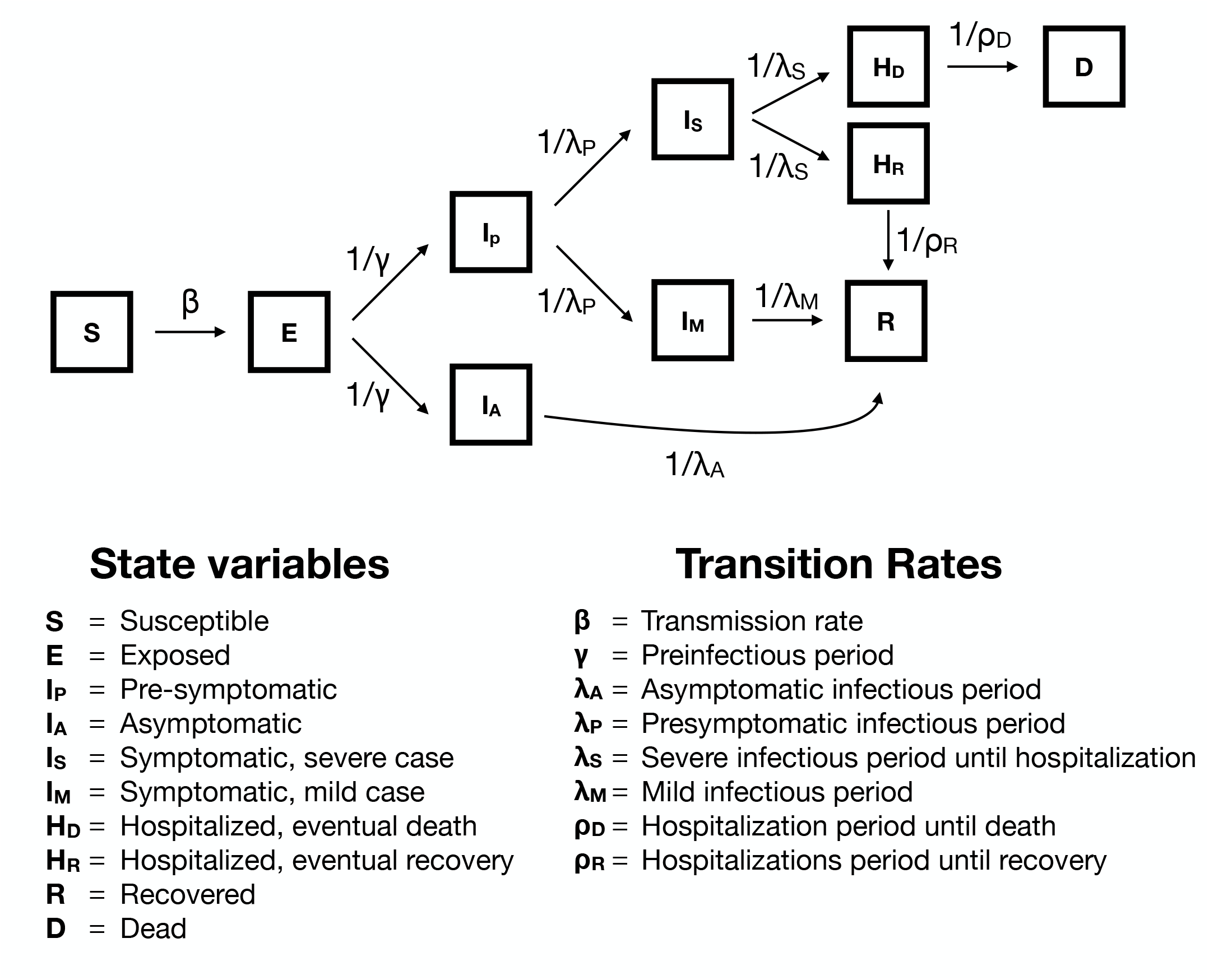
Epidemiological model box diagram

**Figure S2:**
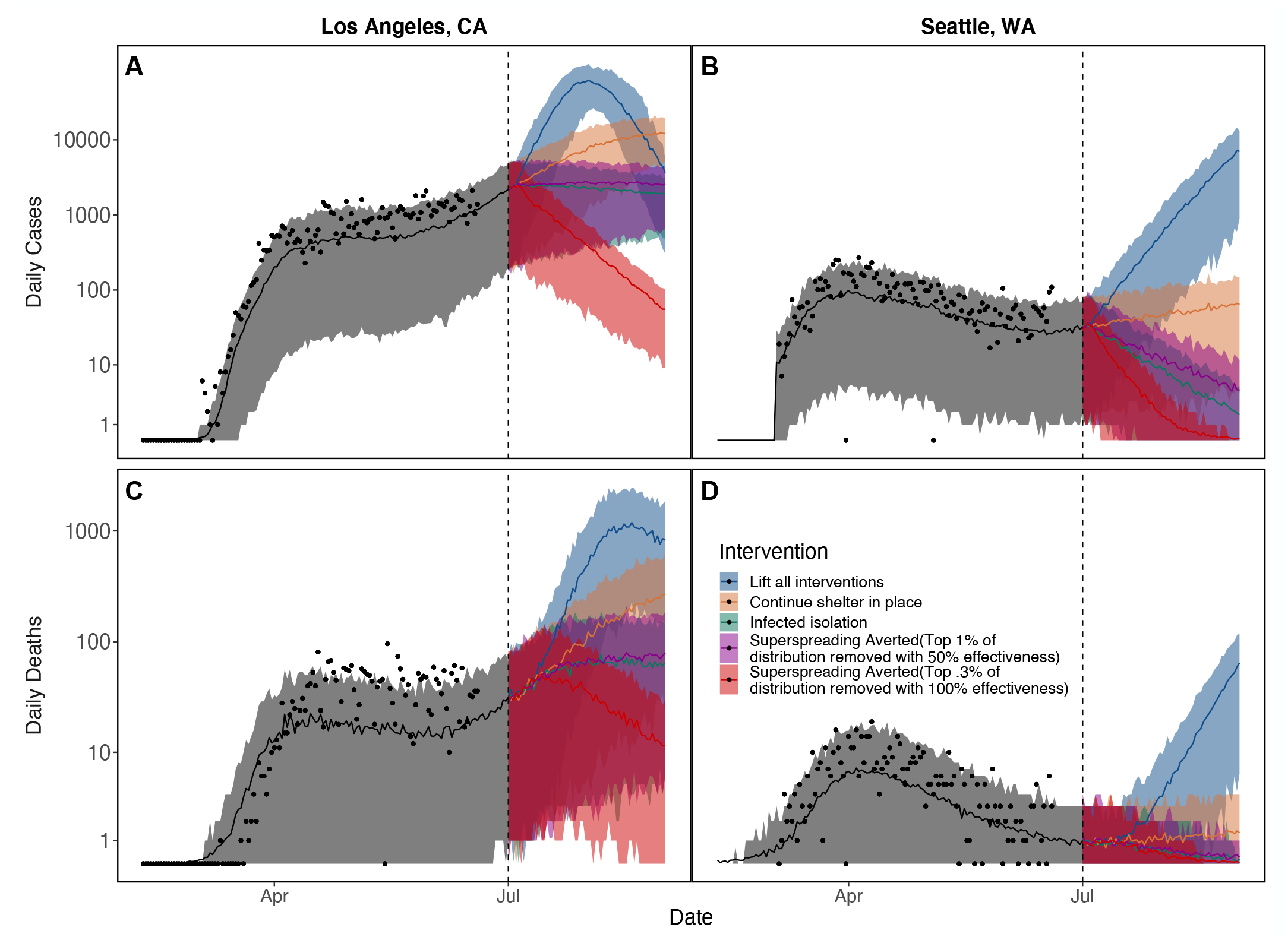
Many other truncation interventions are viable alternatives to the top 0.5% with 75% efficiency presented in the main text including: truncating the top 1% with 50% efficiency (purple) and top 0.3% with 100% efficiency (red). Bands show 95% CI on stochastic simulations of daily cases and deaths for the single maximum likelihood estimate. Dates range from February 2020 to October 2020.

**Figure S3:**
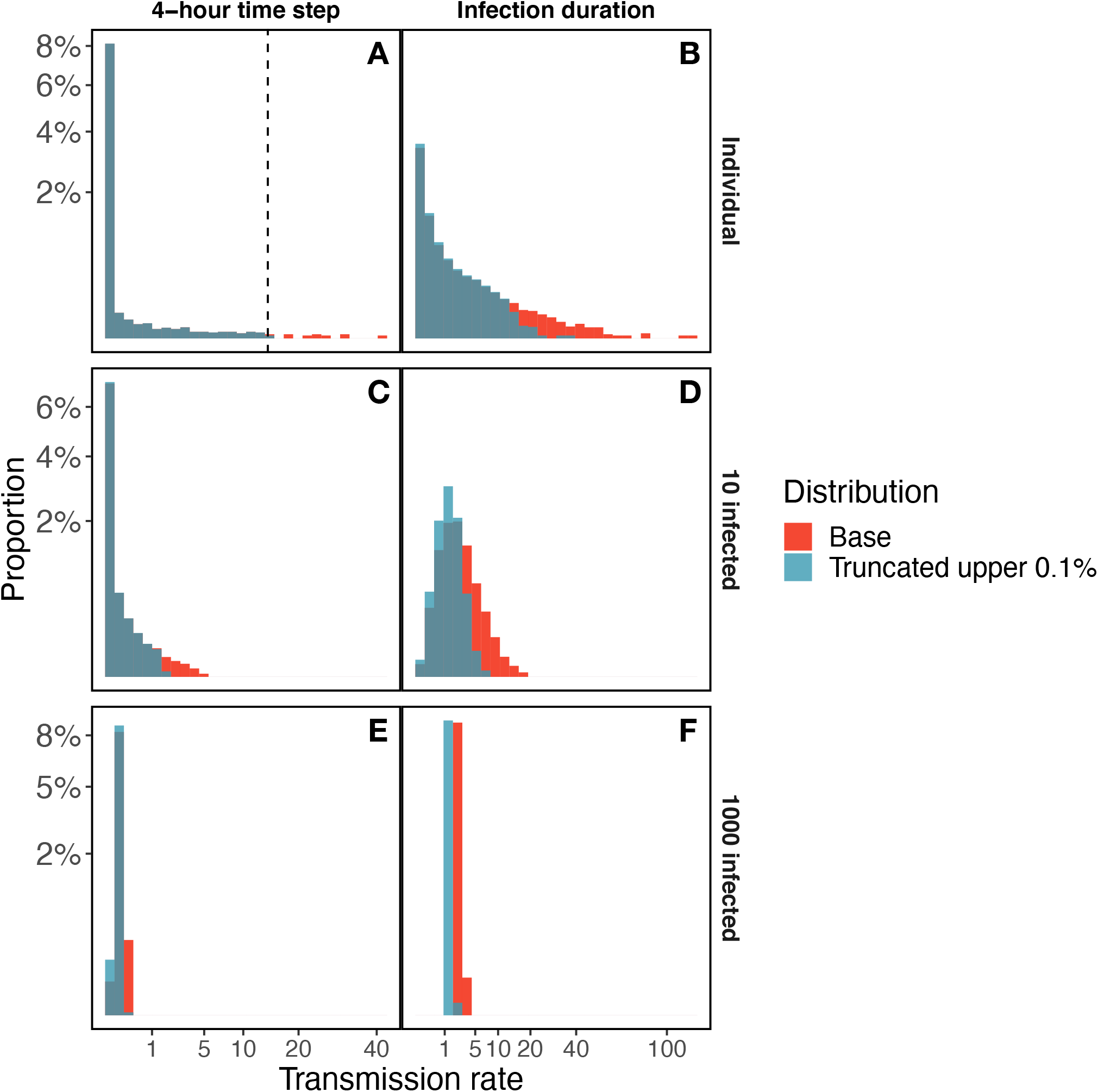
Expanded view of truncating the upper 0.1% of the individual level time step transmission rate distribution (π) at a four-hour time step (A). This truncation leads to a reduction of the mean and variance for an individual’s infectious period reproduction potential (B). As the number of infected individuals in the population increases from 10 (C, D) to 1000 (E, F), the variance in decreases in both the population-level average transmission rate during each 4-hour period (C, E) and over the lifetime of those infected (D, F).

**Figure S4:**
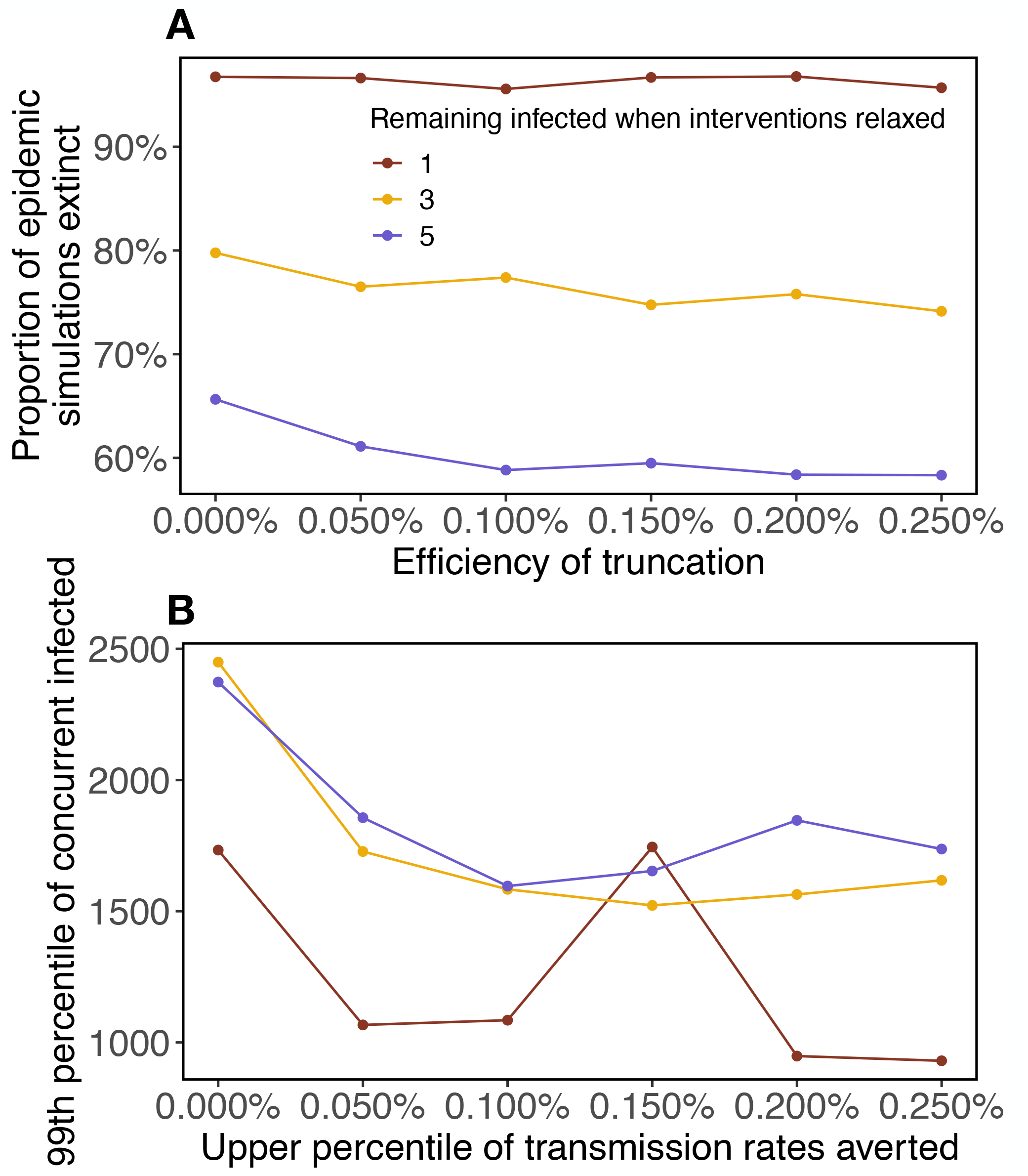
The proportion of epidemic simulations that went extinct (A) and the upper 99th percentile of the number concurrent infected after 42 days (B) for the resurgent simulations among 5000 total simulations for increasing truncation proportions of *π*. Shelter-in-place is scaled so that transmission rate at the time of intervention relaxation is identical across intervention scenarios and would result in ℛ _*E*_ = 2 in a fully susceptible population.

**Figure S5:**
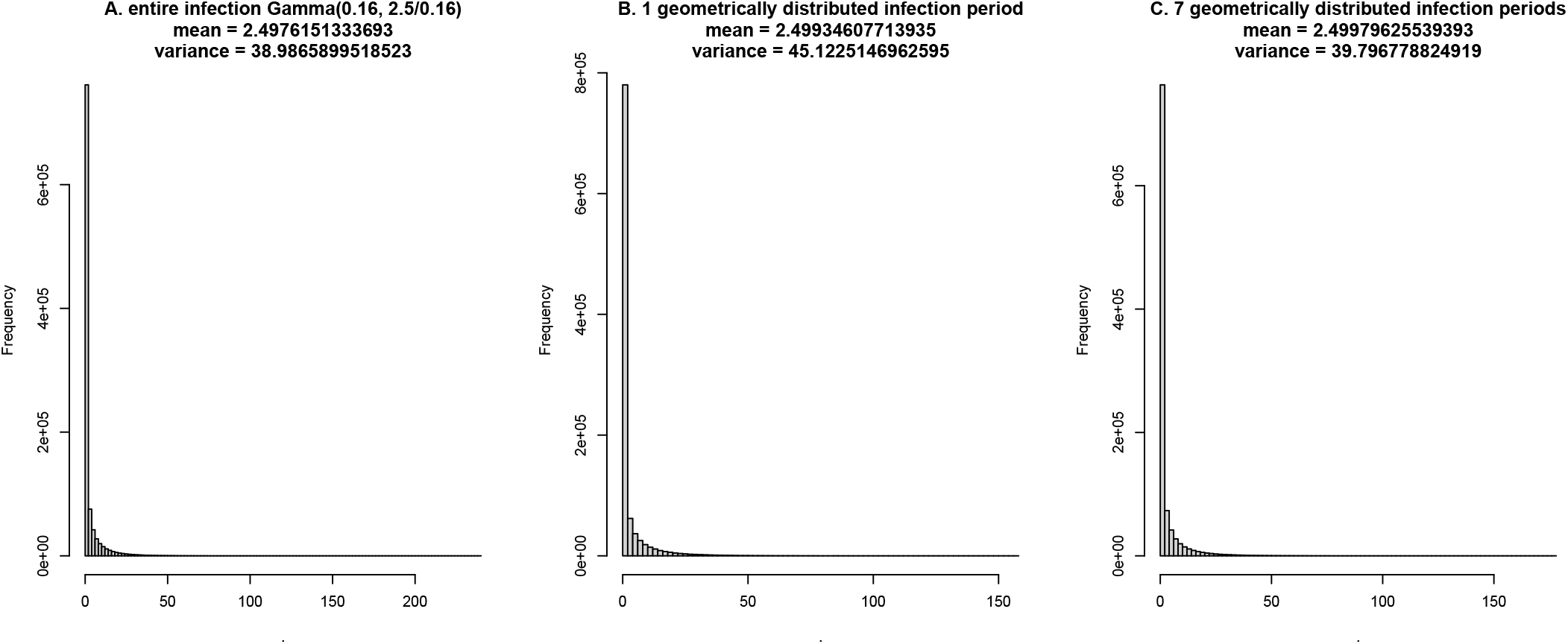
The distribution of individual lifetime reproduction (*R*) when modeled as a Gamma distribution with a mean of 2.5 and a scale of 0.16 (A). This distribution implicitly assumes a constant infectious duration. Using a geometrically distributed infectious period with only one period (“box”), and a time period of 4 hours result in an increase in the variance of the individual reproductive distribution relative to assuming a constant infectious period (B). Breaking the infectious period into 7 sub-stages (boxes) reduces the variance, though the variance remains marginally higher than when assuming a constant infectious period (C).

